# Gut microbiome co-abundance networks varies with age, sex, smoking status and body mass index

**DOI:** 10.1101/2024.04.30.24306630

**Authors:** Christophe Boetto, Violeta Basten Romero, Léo Henches, Arthur Frouin, Antoine Auvergne, Etienne Patin, Marius Bredon, Milieu Intérieur Consortium, Sean P. Kennedy, Darragh Duffy, Lluis Quintana-Murci, Harry Sokol, Hugues Aschard

## Abstract

The gut microbiome is a complex ecosystem whose members develop local interactions to form coherent functional communities. Variability in these communities, typically investigated through taxa co-abundance, might provide critical insights on the biological links between the gut microbiome and human phenotypes. However, existing methods to investigate variations in taxa co-abundance suffer multiple limitations. Here, we address the simple but challenging question of identifying factors associated with variability in gut microbiome taxa co-abundance using a novel covariance-based method that resolve these limitations. We screened 80 host factors in 938 healthy participants, and identified associations between taxa co-abundance variability and age, sex, smoking status, and body mass index (BMI) not captured by abundance-based and diversity-based methods. Increased age and smoking were associated with an overall decrease in co-abundance, and conversely BMI with an increase. Finally, we demonstrate that the proposed approach offers a powerful framework for describing taxa networks at the individual level and predicting host features.

## Introduction

Major initiatives such as the Human Microbiome Project^1^, and epidemiological studies revealed important features associated with changes in the composition of the gut microbiome, including age^2-4^, sex^5-7^, BMI^7-9^, smoking habits^10-12^, long term diet^8,13-17^, and host genetics^17-22^ in both healthy individuals and disease cases^23-25^. Yet, these studies also highlighted important challenges in deciphering the complex host-microbiome relationship^26,27^. In particular, despite its high dimensional nature, most existing human microbiome studies still focus on univariate approaches, testing associations between a variable of interest and each single taxon. To address this limitation, there has been an increasing interest in multivariate approaches. This includes the application of the alpha and beta diversity indexes, which measure intra- and inter-sample distances between microbiome samples^19,22,28^, and various multivariate mean-based approaches to conduct a joint test of the abundance of multiple taxa with a predictor^29-31^. Conversely, the study of features associated with variability in the co-abundance of taxa remains puzzling^32-34^.

The study of gut microbiome taxa co-abundance, which captures the global connectivity within the gut microbiome, is arising as a promising research topic^35-40^. The gut microbiome is a complex ecosystem whose constituents form sub-communities through interactions between individual taxa. Those sub-communities, sometimes referred to as guild^37^, or cliques, display co-abundances because they exploit the same class of resources or work together as a coherent functional group^32,38^. Previous studies already illustrated the potential differences in co-abundance networks across inflammatory bowel disease status and body mass index^35^, and geographically diverse populations^39^. However, there is currently no gold standard method to screen for factors associated with changes in the co-abondance of bacteria across individuals in a population. Existing methods are typically based on the inference and pairwise comparison of networks across conditions, using a threshold-based approach to define co-abundances when the phenomenon is likely continuous. Furthermore, previous works showed that the inferred networks can vary substantially across approaches^41^. More problematic, by construction those methods are restricted to categorical predictors, they do not allow for variable adjustment, and they do not provide a formal global test of association. We recently developed MANOCCA^42^, a formal statistical framework to test the effect of both categorical and continuous predictors on the covariance matrix of a multivariate outcome –a metric directly proportional to the co-abundance– that addresses these methodological gap.

Here, we applied MANOCCA to study host factors associated with variability in the gut microbiome co-abundance network of 938 healthy participants from the Milieu Interieur cohort^43^, and compared results with multiple standard univariate and multivariate approaches. We then conducted an in-depth examination of the effect of each associated factor on the taxa interaction network, highlighting the key taxa impacted and how these factors shape the microbiome composition. Finally, we used our framework to assess the performances of predictive models of the associated factor based on taxa co-abundances.

## Results

### Identifying host factors associated with taxa co-abundance

The composition of the gut microbiome of a single individual can vary with the many host factors driving the molecular environment of the gut. Exposure to a given environment and host characteristics might promote specific mechanisms, inducing a collaboration across a limited number of species, while another environment would promote only a subset of the same species. The overlap of such mechanisms can result in various co-abundances independently of a mean effect of the exposures in question (**Fig. 1**). We screened 80 host variables, including demographics, socio-professional, and dietary habits measurements (**Table S1**), for association with overall taxa co-abundance network in healthy individuals using MANOCCA. We conducted analyses at the species, genus, and family levels, focusing on the most common taxa^44^, including respectively 675, 718, and 151 taxa after quality control filtering (**Fig. 2** and **Table S2**). Except when used as predictors, all analyses were adjusted for age, sex and body mass index (BMI).

**Figure 1.**
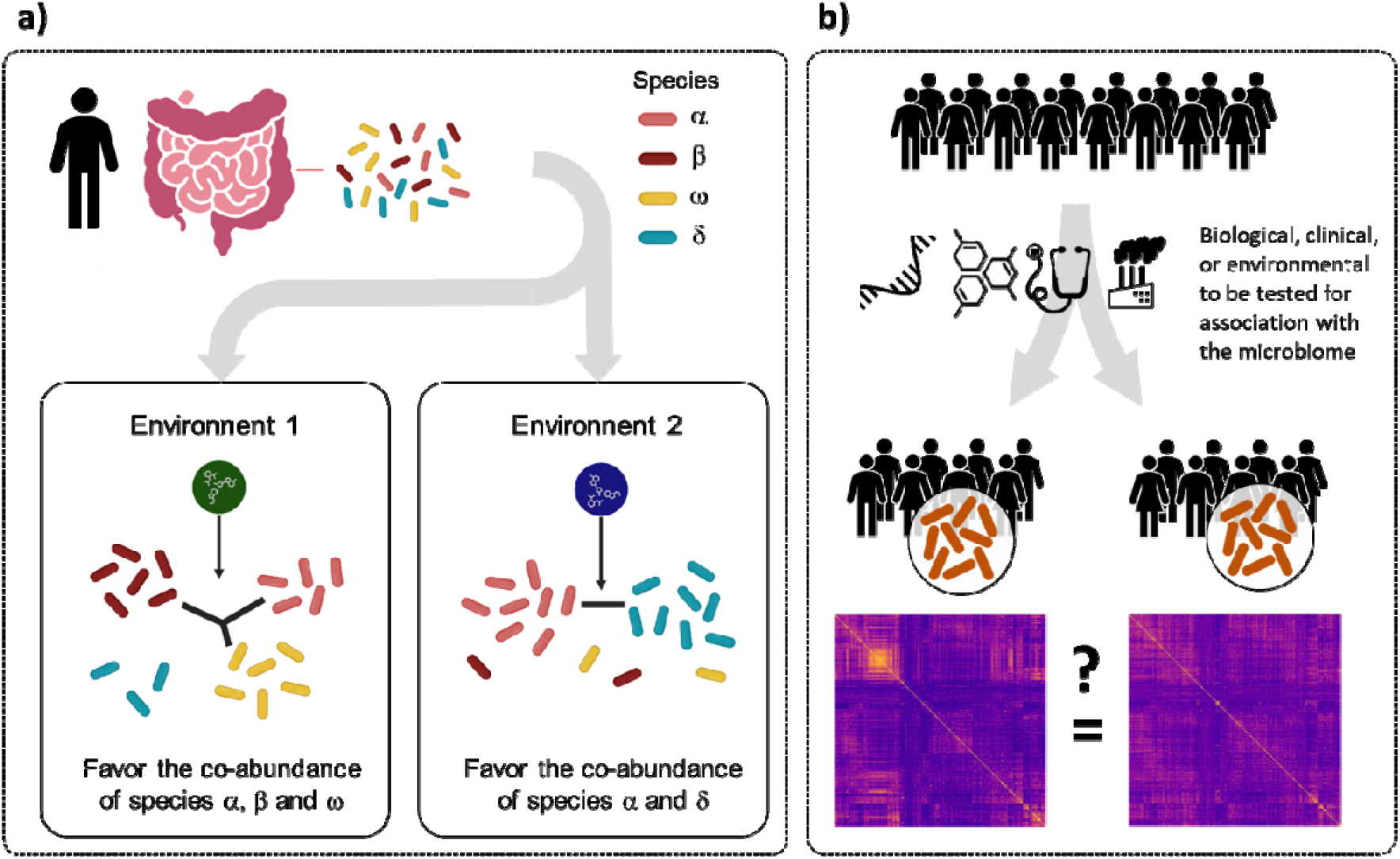
Co-abundance testing principle. Panel a) illustrates the targeted mechanism at the individual level. The equilibrium of a given microbiota ecosystem can change conditionally on the environment, impacting not only the marginal abundance, but also the co-abundance of species. Under environment E1, the available local resources foster a complementary system involving species α, β, and ω. Under environment E2, the available local resources foster a complementary system involving species β, and δ. Panel b) illustrates an application scenario at the population level, where health features (biological, clinical or environmental variables) are tested for association with the covariance matrix of the microbiome measured in the same individuals. When the feature of interest is binary (e.g. exposed vs unexposed), the approach consists in testing for statistical differences in the observed microbiome covariance derived in exposed and unexposed participants separately. When the feature of interest is continuous, the same principle applies.

**Figure 2.**
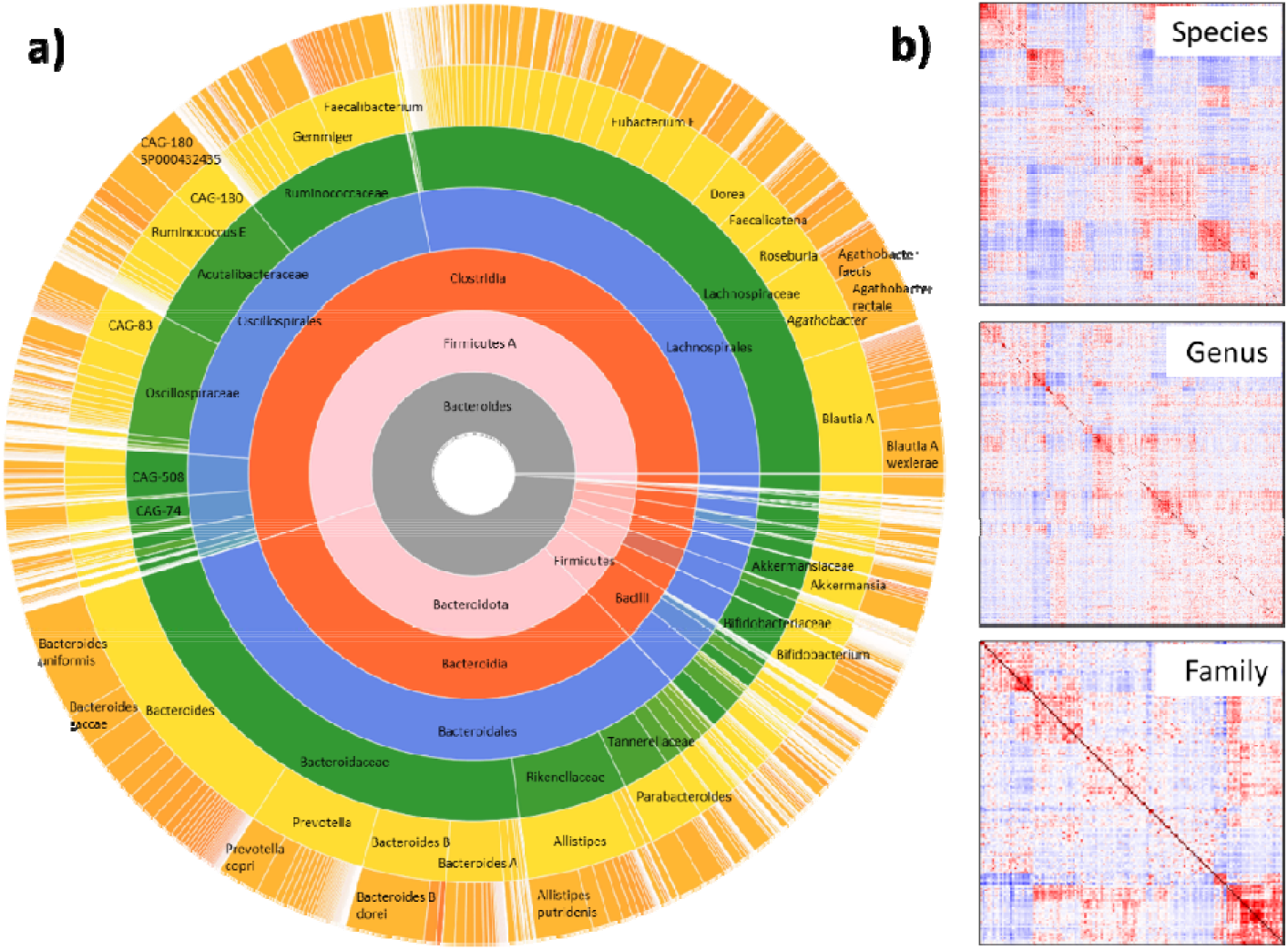
Milieu Interieur microbiota composition. Panel a) shows the relative abundance of taxa across the seven taxonomic levels (Kingdom, Phylum, Class, Order, Family, Genus, Species). Panel b) shows the covariance matrices at the Family, Genus, and Species levels derived from all 938 patients.

We identified associations with co-abundance variability at all three taxonomic levels significant after correction for multiple testing (*n*= 8,000 tests, *P* threshold = 6.25 x 10^-6^) with age (*P*_*species*_ = 2.0 x 10^-55^, *P*_*genus*_ = 3.5 x 10^-56^, *P*_*family*_ = 9.2 x 10^-37^), sex (*P*_*species*_ = 2.2 x 10^-17^, *P*_*genus*_ = 6.3 x 10^-22^, *P*_*family*_ = 3.1x 10^-18^), smoking (*P*_*species*_ = 2.8 x 10^-14^, *P*_*genus*_ = 1.6 x 10^-20^, *P*_*family*_ = 5.6 x 10^-13^) (**Fig. 3**). Analyses at the genus level further identified an association with body mass index (*P*_*genus*_ = 5.9 x 10^-6^). Note that MANOCCA uses a dimension-reduction step to address the large number of parameters in the taxa covariance matrix. Varying the parametrization of this step did not qualitatively impact the results. Details of this analysis are provided in **Figures S1-S2** and **Supplementary Notes**. We also conducted sensitivity analyses, assessing the variability in the results when applying MANOCCA to random subsets of available taxa. Overall, the larger the set of taxa included, the stronger the association signal (**Fig. S3**), highlighting a global effect of the four factors on the microbiome co-abundance network.

**Figure 3.**
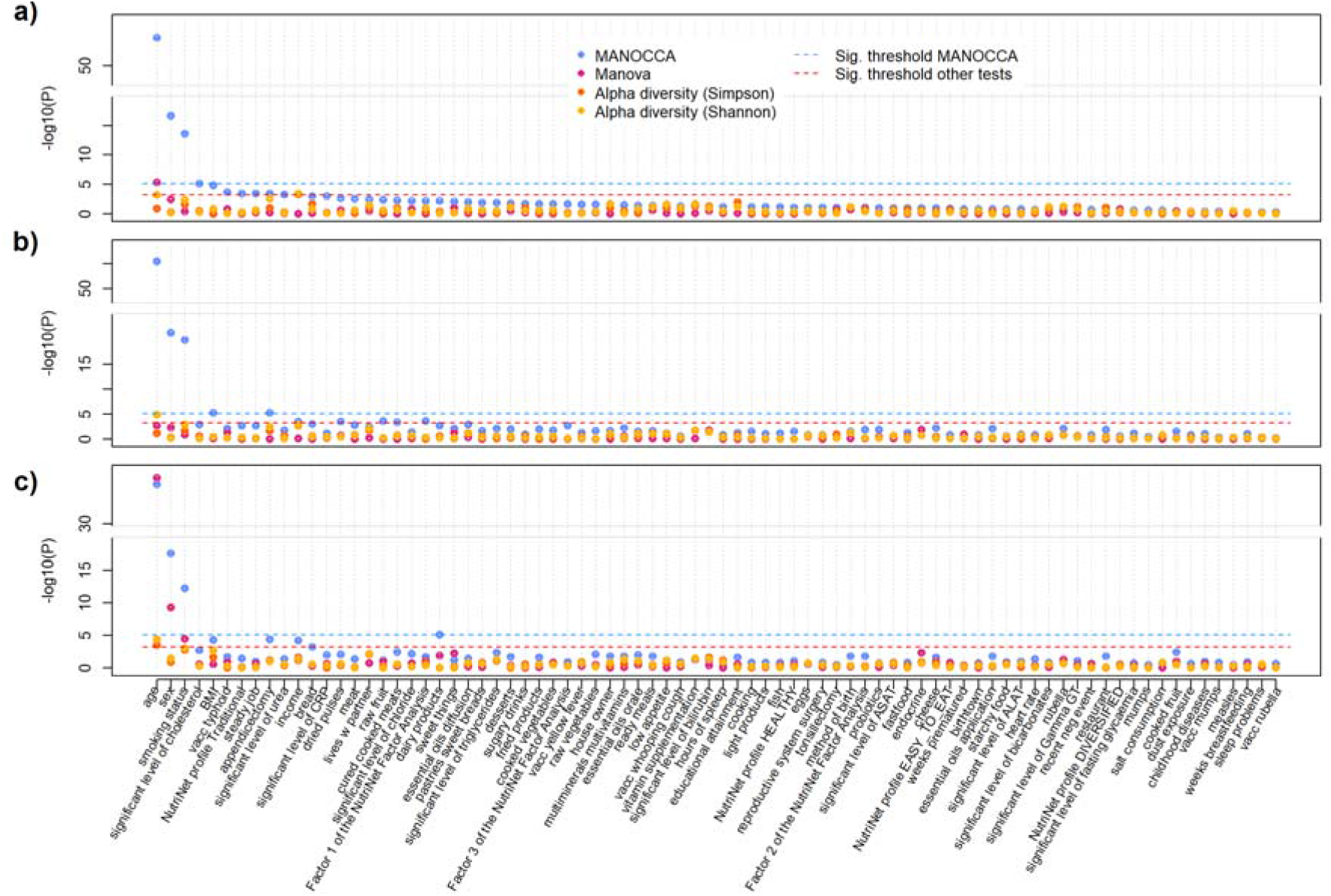
Screening for environmental and clinical factors. Association screening between 80 environmental and clinical factors from the Milieu Interieur cohort and the covariance of taxa at the species (a), genus (b) and family (c) taxonomic levels. Each panel display the -log10(_P_) of each predictor. Results from MANOCCA are based on the optimal number of principal components. Results from MANOCCA are compared against three alternative approaches: a standard MANOVA, and alpha diversity tests based on both Simpson and Shannon metrics. The red dash line indicates the stringent Bonferroni correction threshold accounting for all predictors and sets of PCs tested.

We applied two alternative multivariate approaches for comparison purposes: a standard MANOVA, testing for association between each host factor and the joint abundance of taxa, and alpha diversity using the Shannon and Simpson indexes (**Fig. 3, Table S3**). Some factors were significant after correction for multiple testing (*P* threshold of 6.3 x 10^-4^ accounting for the 80 tests per approach) but at a much lower significance level. The MANOVA identified an association with age at the species and family level (*P*_*species*_ = 5.2 x 10^-6^, *P*_*family*_ = 1.0 x 10^-29^) and with sex at the family level (*P* = 5.2 x 10^-10^). Both Simpson and Shannon indexes identified a signal with age at the family level (*P*_*Simpson*_ = 3.4 x 10^-4^, *P*_*Shannon*_ = 3.9 x 10^-5^) and the Shannon index also identified age at the genus level (*P*_*Shannon*_ = 1.6 x 10^-5^). Altogether, the additional signals observed with smoking and BMI, and the stronger association of age and sex with the covariance-based approach as compared to mean-based and diversity index approaches, points towards a substantially larger information content of the co-abundances of taxa over these existing metrics to describe the relationship between the gut microbiome of healthy individuals and these host variables.

### Contribution of taxa on the co-abundance association signal

By construction, the MANOCCA statistic is a weighted sum of the contribution from each pair of taxa (see **Methods**), so that the contribution weight of each taxon on the co-abundance association signal can be easily extracted. We derived these weights for the age, sex, smoking and BMI signals, focusing on the genus level, which displayed the strongest association. As shown in **Figure S4a**, most taxa display a non-zero contribution to the association, highlighting again the global effect of these factors on the microbiome composition. Nevertheless, these contributions show substantial heterogeneity, with a limited number of taxa pairs displaying outstanding weights as compared to the expected under the null (**Fig. S4a-b**). To assess potential links between effects on co-abundance and effects on relative abundance, we compared the MANOCCA weights against the univariate mean effect *P*-value associations derived using a linear regression (see **Methods** and **Table S4**). As shown in **Figure S4c**, we observed a positive and significant correlation between the two terms for all four variables (age, *P* = 1.6 x 10^-12^; sex, *P* = 1.3 x 10^-40^; smoking, *P* = 2.1 x 10^-5^; BMI, *P* = 4.6 x 10^-15^), suggesting a dual impact of these factors on the abundance and the co-abundance of many of these genera, in agreement with the existing literature^2-12^. The correspondence is especially marked for sex. Several top genera contributing to the co-abundance also pass a stringent Bonferroni correction threshold (*P* < 8.7 x 10^-7^) for univariate association. This includes *Bacteroides* (*P* = 1.9 x 10^-7^), *Coprococcus* B (*P* = 7.1 x 10^-9^), *Anaerotruncus* (*P* = 8.5 x 10^-7^), *Agathobacter* (*P* = 3.0 x 10^-8^), *Alistipes* (*P* = 2.2 x 10^-8^), and *Intestinimonas* (*P* = 1.9 x 10^-9^).

We next investigated the characteristics of the top 5% pairs of genera displaying the largest contribution to the variability in co-abundance at the family level. Out of 151 families, a subset of 9, 7, 9 and 6 overlapping sets of families, covered 50% or more of those top contributing genera with age, sex, smoking and BMI, respectively. Those key families include the ones with the highest relative abundance, *Lachnospiraceae* (23.6%), *Bacteroidaceae* (22.4%), *Ruminococcaceae* (7.8%), *Acutalibacteraceae* (6.4%), *Oscillospiraceae* (5.6%), but also some rare ones: *Eggerthellaceae* (0.5%), *Peptostreptococcaceae* (0.6%), *Muribaculaceae* (0.5%), and four unspecified Co-Abundance Groups (*CAG-74, CAG-508, CAG-272, CAG-138*) (**Fig. 4a**). While the representativity of families involved in co-abundance variability was somewhat proportional to their relative abundance, we noted several major differences. Some families, such as *Bacteroidaceae* are largely underrepresented in the co-abundance signal. Conversely, co-abundance involving the *Oscillospiraceae* family are strongly impacted by all four factors, and by BMI in particular. Other families also display factor-specific enrichment, including *Peptostreptococcaceae* and *Muribaculaceae* with smoking, two families already reported to be associated with smoking status^45,46^.

**Figure 4.**
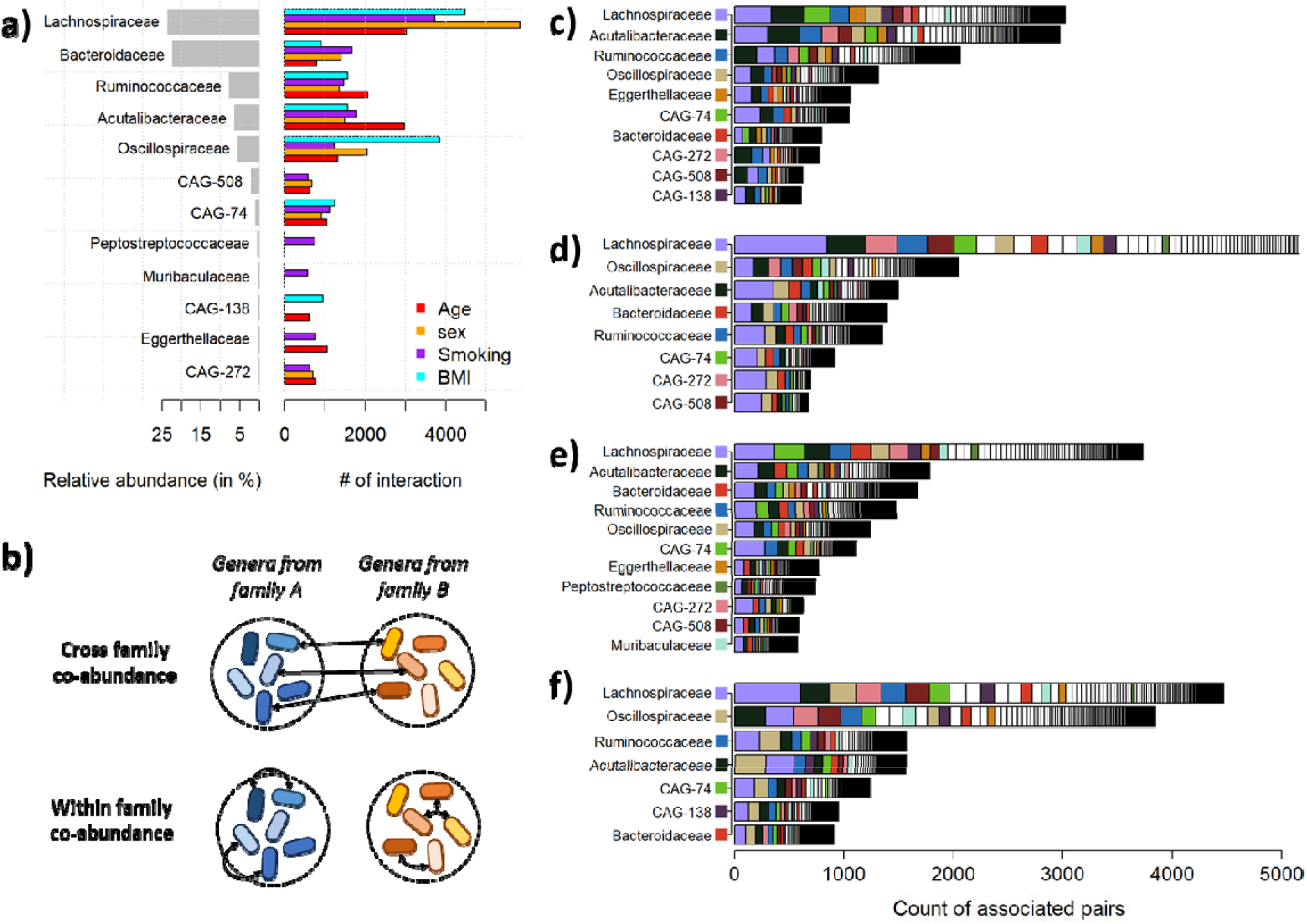
Cross and inter-family interactions for age, sex, smoking and BMI. We used the top 5% pairs of genera with the largest contribution to the covariance test to investigate whether variability in co-abundance involved genera from the same family or from different families. Panel (a) presents the relative abundance of the family from these top genera in the whole cohort (grey bar), and the number of interactions observed for genera from each of these families. Panel (b) illustrates the difference between cross-family and within family interactions. Detailed co-abundance pairs are presented for age(b), sex(c), smoking(d) and BMI(e). The Y axis represents the families of the top subset of genera involved, and that together explain up to 50% of the signal. The X axis shows the distribution of families for the associated genera, defined as a count per family. The length of the bar indicates how many pairs are involved for each top family. Colours were assigned only for the top families, and white blocs correspond to the unlisted categories.

Finally, we examined the composition of the top contributing pairs of genera to determine whether they involve changes in interaction within the same family (intra-family co-abundance) or interaction between genera from different families (cross-family co-abundance) (**Fig. 4b**). Within-family co-abundance represented a small fraction of all interactions, with the vast majority of interactions taking place between genera of different families (**Fig. 4c-f**). Besides a few exceptions (e.g. variability in the co-abundance between taxa from *Oscillospiraceae* and *Acutalibacteraceae* families for BMI), we did not observe any marked pattern.

### Network of impacted taxa

We formed a network of co-abundance variation from the top 1,000 pairs of genera contributing to the MANOCCA association signal with age, sex, smoking, and BMI (**Fig. 5a**, and **supplementary Material**). Altogether, these 4,000 pairs involved a total of 476 unique genera. As shown in **Figure 5b**, there was a substantial overlap in pairs of co-abundant taxa impacted by sex and BMI (N= 658 pairs, approximately 66% of sex and BMI associated pairs), and age and smoking habits (N=306 pairs, approximately 31% of age and smoking-associated pairs). Conversely, the overlap across the nine other pairs of factors was null or negligible. At the taxa level, a core of 200 genera were shared across all predictors (**Fig. 5c**). Other genera were evenly spread across factors, except for age and smoking which involved 49 (13%) and 54 (15%) genera specific to those two factors, respectively. Together, this suggests that the four factors partly control the interacting partners of this core genera. As shown in **Figure 5d**, increased age and smoking are mostly associated with a decrease in co-abundances, with 86% and 75% of top pairs displaying negative associations with these two factors, respectively. BMI exhibited an opposing trend, with 72% of top pairs showing an increase in co-abundance with increasing BMI. The sex predictor displayed a more balanced distribution, with a 60% decrease and a 40% increase of co-abundance in males as compared to females.

**Figure 5.**
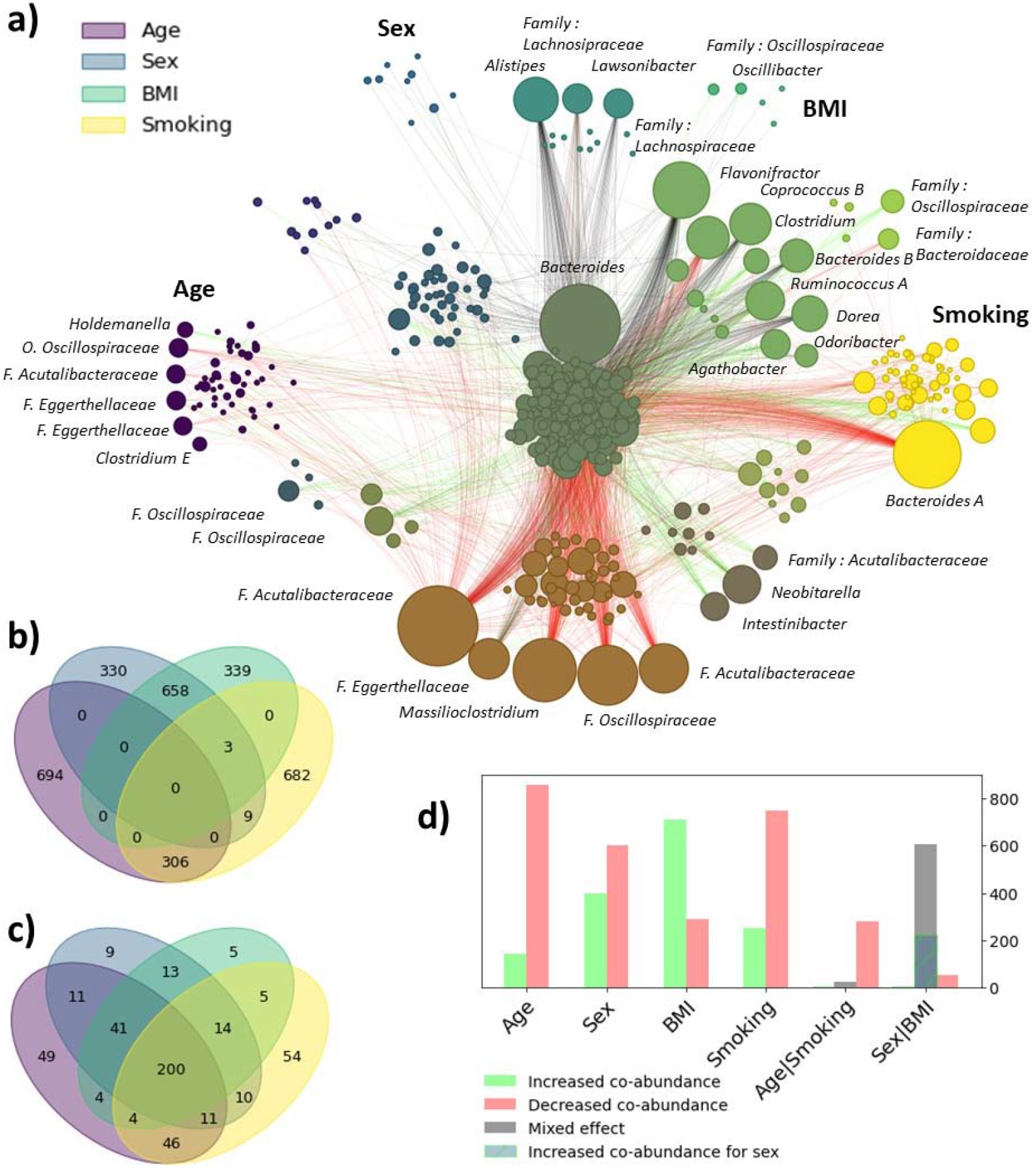
Co-abundance network influenced by age, sex, smoking and BMI. For the top four associated features from the MANOCCA (age, sex, BMI and smoking), we extracted the top 1000 contributing pairs of genera out of the 259,560 total products and derived the direction of effect of each predictor on the pair of co-abundance. We plotted the Venn diagram of shared pairs between each feature in a) and the overlap in taxa in b). In c) we show the distribution of direction of effects per predictor, and for the age – smoking and sex – BMI intersections. We then used the pairs of features to derive a network of the changes in correlation with regard to each predictor. The node size, representing a genus, is proportional to its number of contributions with other genera, and edges link the top contributing pairs. The edge colors indicates the direction of effect with green indicating that an increase of the predictor drives an increase in co-abundance, red shows that an increase of the predictor drives a reduction in co-abundance and black indicates a mixed direction of effect for the overlapping predictors. The color of each node depends on how it is shared across the four predictors, and follows the structure of the b)-c) venn diagrams.

Multiple patterns emerged when exploring the contributing genera. Those shared between smoking and age are especially enriched in the *Oscillospirales* order (e.g. *Massillioclostridium, CAG-180, CAG-1427, Marseille-P4683, and MGYG-HGUT-03297*), and consistently exhibited reduced co-abundances with the core taxa. Among genera unique to smoking, *Bacteroides A* genus was by far the most impacted, showing a reduction of co-abundances with many of the core taxa. Interestingly, the relative abundance of this common genus (detected in 99% of participants, **Table S2**) was not associated with smoking status in our data (*P*-value from a linear regression equals 0.26, **Table S4**). This suggests that smoking might only break some of its interactions with other genera without impacting the presence of this genus itself, highlighting the ability of our approach to detect taxa missed by standard abundance-based approaches. A subset of genera contributing to association with BMI, sex and smoking was enriched from the *Lachnospiraceae* family (*Ruminococcus A, Dorea, Coprococcus B, GCA-900066135, Agathobacter*), displaying both increased and decreased co-abundances across predictors. Both *Lachnospiraceae* relative abundance and co-abundance with other taxa have already been found to be associated with human diseases and obesity in particular^35,47,48^.

Finally, to assess the relevance of the covariance-based co-abundances network impacted, we considered two alternative approaches. A naïve permutation-based approach, inspired from the existent^41^, that produces an empirical comparison of pairwise covariance between all taxa (see **Methods**), and the commonly used SparCC^49^ approach (see **Supplementary Notes** for a detailed description of the two approaches). Note that both the SparCC and the permutation-based approaches, like all existing method, are limited to binary predictors and use a threshold to define co-abundance in each group studied. Both methods are meant to detect significant differences in pairwise taxa correlations across values of a categorical predictor, and should in theory detect effect on co-abundance variability similar to those detected by MANOCCA. However, as showed in the simulation from **Figure S5**, permutation-based shows poor specificity as compared to MANOCCA, and SparCC shows the poorest performances in this simulation with almost no power. We applied both methods to the two binary predictors, sex and smoking, at the genus level and crossed the results with MANOCCA’s top contributing products. The overlap between MANOCA and the two alternative methods was very modest, but highly significant with minimum *P*-values of 1 x 10^-145^ and 2 x 10^-153^ for sex and smoking respectively for the permutation approach (**Fig. S6 a-b**), and 1 x 10^-70^ and 1 x 10^-82^ for sex and smoking respectively for the SparCC approach (**Fig. S6c-d**), thus, confirming that those three alternative methods do detect some similar network components.

### Prediction of individual features based on taxon correlation

Our framework is built out of a linear model where the covariance is defined at the individual level. This is a major advantage over existing correlation approaches^41^, that allows for a range of complementary analyses. One particularly important extension is the possibility of training a predictive model of an outcome based on taxa covariance, so that the outcome in question can be predicted for any new individual based on its microbiome (and conversely). Here, we assessed the accuracy of MANOCCA to predict the four most associated features (age, sex, smoking and BMI), using taxa from the species, genus and family level and a 30-fold cross-validation. Accuracy was derived using squared-correlation (*r*^2^) for continuous outcomes (age and BMI), and using the area under the receiver operating curve (AUC) for binary outcomes (smoking and sex). We compared the covariance-based prediction model against a standard linear model based on the relative abundance of each single taxa.

As showed in **Figure 6**, the MANOCCA strongly outperforms the standard mean-based prediction model, being significantly more accurate in all scenarios we considered. Gain in prediction was especially large for age with up to a three-fold increase in power. The median of 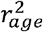 from the MANOCCA equals 0.27, 0.25 and 0.18 for models based on species, genus and family, respectively. In comparison, the mean-based model 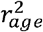 equal 0.10, 0.07 and 0.05, respectively. Prediction was also significantly higher for sex, with AUCs of 0.66, 0.64, and 0.64 for the mean-based model at the species, genus and family level, respectively, and AUCs of 0.67, 0.69 and 0.70 for the covariance-based model. This confirms the higher information content of co-abundance as compared to abundance, and demonstrates the validity of using covariance-based co-abundances for prediction purposes.

**Figure 6.**
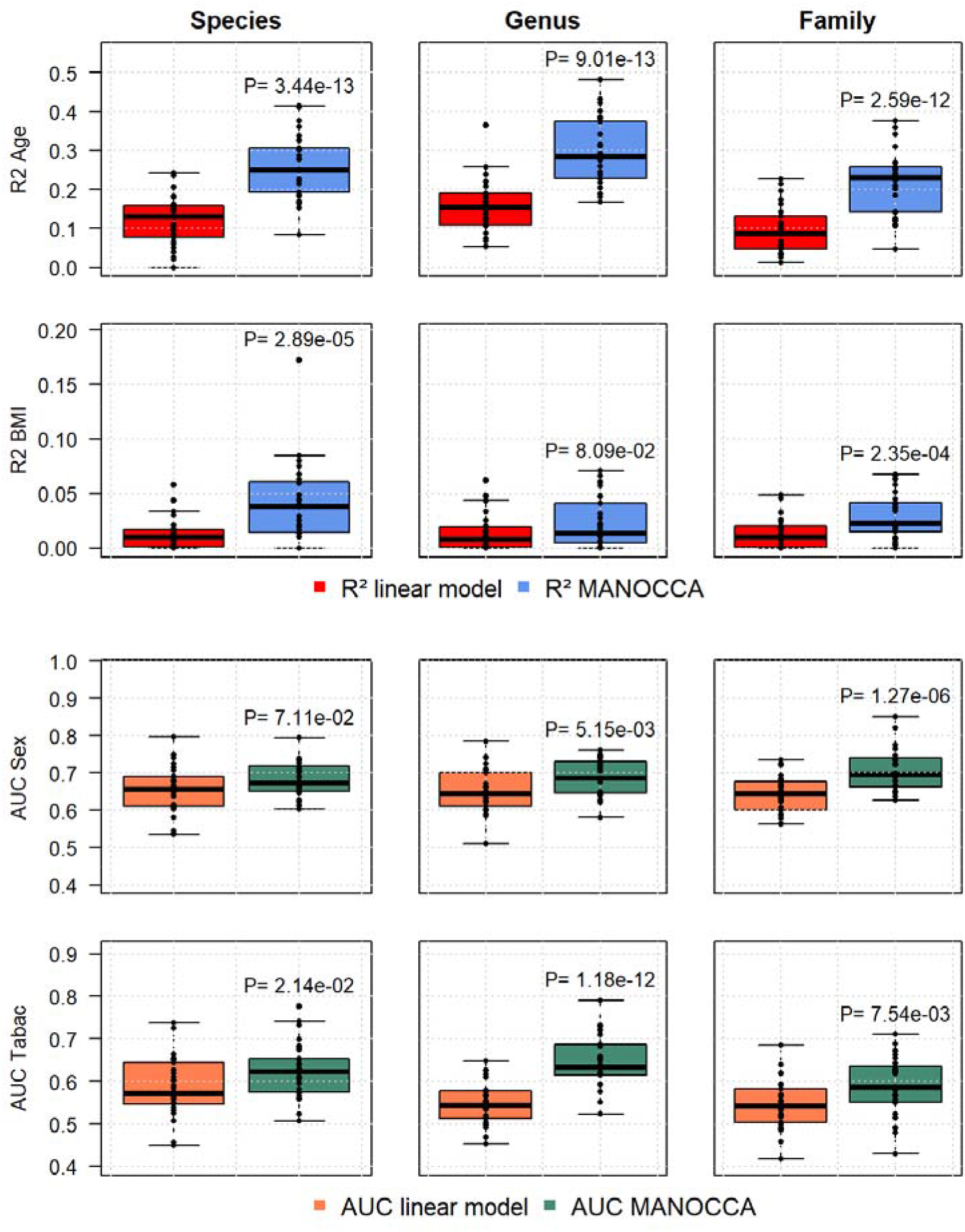
Predictive power of covariance-based models. We estimated the predictive power of co-abundance models (blue/green boxplots) as compared to the standard mean-based multivariate model (red/orange boxplots). For each of the four most associated features (age, sex, smoking and BMI) and the three lower taxonomic levels (species, genus, family), we derived the prediction accuracy using the squared-correlation (R^2^) for continuous features and the area under the receiver operating curve (AUC) for binary feature. The analysis was done using a 30-fold cross validation, with the score being train in 90% of the data, and the R2 being derived in the remaining 10%. The significance of the difference between the two models was tested using a two-sample two-sided t-test.

## Discussion

There is a strong rationale for studying microbiome co-abundances. There is now increasing evidence that species form functionally coherent groups that work together to exploit the same resources from the local environment^38^. Studying those groups, rather than each single taxa, might help better understand the role of the microbiome in human health outcomes. With this same argument, it has already been proposed to study those groups through variability in the network of co-abundances^35,39^. Although simple in principle the implementation of this objective can be challenging in practice. Here, we applied MANOCCA^42^, a recently developed method, that allows to conduct a formal statistical test of association between taxa covariance and any predictor, whether continuous, categorical or binary, to investigate host features associated with the gut microbiome co-abundance in 938 healthy individuals from the Milieu Interieur cohort.

We identified highly significant associations between taxa co-abundance variability and age, sex, smoking status and body mass index (BMI). Except for BMI, these associations were detected at all three taxonomic levels studied: species, genus, and family. In comparison mean-based multivariate and diversity-based analyses only identified associations with age, and one association with sex at the family level, but at a much lower significance level. For the four associated features, there was a significant correlation between contribution to co-abundance and univariate effect on relative abundance, suggesting that those features impact both the abundance and co-abundance of taxa. The network of top contributing genera shows that variability in interactions were concentrated in a limited number of families and are essentially taking place between genera of different families, rather than between genera of the same family. The overlap of top contributing taxa over the four features was substantial, especially between age and smoking, and between sex and BMI, suggesting potentially shared mechanisms. Finally, we demonstrate that the MANOCCA framework can be used to build predictive models. In this study we applied it to age, sex, smoking and BMI. For all features, the predictive power based on co-abundances was significantly and systematically higher than for a standard mean-based multivariate model, with up to a three-fold increase in r-squared for age.

Our study also has limitations. First, the approach is not applicable to microbiome data of small sample sizes. Despite the data reduction steps through principal component analysis, the number of PCs analysed should remain substantially larger than the sample size, thus limiting the application to datasets of 100 participants or more. Hopefully, this will become less of an issue thanks to the increasingly large cohorts available. Second, because of that data reduction step, each application requires the selection of a number of principal components to be kept. While the optimal parameters are likely to change across data, this analysis suggests that the gain of a systematic screening over a range of PCs can overcome the cost of additional multiple testing corrections. Third, the proposed approach does not model the compositional aspect of the data per se^50^. However, when the dimension of the data is large enough, as for the analysis of species, genus or family, this issue becomes negligible (**Supplementary Notes** and **Fig. S7**). More importantly, the proposed approach assess variability in the covariance, and under reasonable assumptions, this variability is independent of the absolute correlation, so that any remaining bias due to the compositional aspect acts as an offset without impacting our test. Fourth, we demonstrated that covariance can be used for prediction purposes, however, the implementation of such predictive model will have to be explored further. As for prediction model based on relative abundance, some species might not be quantified in the targeted samples for prediction. This issue will be exacerbated when working with thousands of covariance terms. One possible solution is to develop sparse predictive models focusing on pairs of taxa that are fairly common, instead of using the entire covariance matrix. Furthermore, we used simple linear predictive models for both abundance and co-abundance. Future work might investigate the use of more complex methods^51^ to combine the proposed covariance into prediction models.

Through the characterization of the links between variability in gut microbiome taxa co-abundance and healthy individual host factors, this study addresses three major limitations of the existent. First, the proposed approach allows for a formal statistical test of association between taxa co-abundance and both binary and continuous host features when all existing methods are restricted to *ad hoc* comparisons of inferred networks across a limited number of conditions. Second, our framework allows for covariate adjustment, so that the respective effects of correlated factors can be deciphered from one another. Third, our covariance-based approach provides a mean to derive a co-abundance metric at the individual level, allowing for a range of secondary analyses, including the development of co-abundance-based predictive models. Altogether, the proposed approach open paths for various co-abundance analyses. It is highly complementary to recent efforts to develop experimental design to study co-abundance (e.g. ^52^). It can be used to produce new working hypothesis, and assess statistical evidence for effect on co-abundance from both observational and experimental data.

## Methods

### Milieu Interieur gut microbiome data

The Milieu Intérieur Consortium is a population-based cohort initiated in September, 2012^43^. It comprises 1,000 healthy volunteers, all recruited in the suburban Rennes area (Ille-et-Vilaine, Bretagne, France), with a 1:1 sex ratio (500 males, 500 females) and an equal distribution across 5 decades (20 to <30 y, 30 to <40 y, 40 to <50 y, 50 to <60 y). The primary objectives of the MI Consortium are to define the naturally occurring variability in a healthy population’s immune phenotypes and to characterize genetic, environmental and clinical factors driving this variability. The cohort collected a broad range of variables, including genetic, genomic, and environmental data, on most participants. On their first visit the volunteers were also asked to fill in an extended form about socio-demographic, lifestyle and family health history, all recorded in an electronic case report form (eCRF). Gut microbiota composition was obtained from shotgun metagenomics sequencing, and taxonomic levels were reconstructed by summing the normalized abundances within a branch at a given level (**Fig. 2**), resulting in a total of 13,446 unique bacterial species. Further description of the data generation are provided in **Supplementary Notes**.

### Covariance method

Variability in the correlation between two standardized outcomes *Y*_1_ and *Y*_2_ can be investigated through the element-wise product of those outcomes. The Pearson correlation coefficient between *Y*_1_ and *Y*_2_ is expressed as 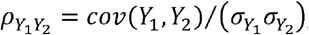 with *cov*(*Y*_1_, *Y*_2_) =𝔼 [*Y*_1_, *Y*_2_] -𝔼 [*Y*_1_] 𝔼 [*Y*_2_] . For standardized outcomes and a sample size *N*, it can be re-expressed as the average of the element-wise product across individuals: 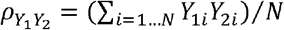 . It follows that the effect of a predictor *X* on *cor*(*Y*_1_, *Y*_2_) can be tested using a standard least-squares regression framework where *X* is treated as a predictor and the product *Y*_1_ *Y*_2_ as the outcome. One can easily demonstrate that, under reasonable assumptions, this test is independent of mean and variance effect^42^.

Extending the method to more than two outcomes can be done through the following three steps: i) starting with *K* centered outcomes *Y*_1_,…, *Y*_*K*_, all the pairwise products are computed: *P*_*ij*_ = *Y*_*i*_*Y*_*j*_ for *i* ∈ ⟦1, *K*⟧ *and i*<*j;* ii) The *P*_*ij*_ products are then mapped to the quantiles of a normal distribution using an inverse-rank normal transformation ; iii) To reduce the dimension of product matrix, *P* is then projected in a reduced latent space of dimension 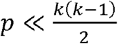 using the Principal Components Analysis transformation: 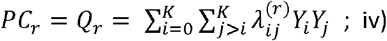 The resulting Principal Components (PC) are then mapped to the quantiles of a normal distribution using an inverse-rank transformation, and scaled. This gives, for *N* considered individuals, a matrix **Q** of dimension *N* × *p* that we can use for the test. A detailed explanation of each step is available in^42^. Finally, given a scaled predictor *X* and scaled covariates matrix **C**, which can be continuous, categorical or binary, the test for association between the predictor and the covariance matrix can be conducted using a standard multivariate model: **Q**∼*X* + **C**.

When applied to taxa, we varied the number of principal components used in MANOCCA from two to one hundred but limited the number of PC analysed for each predictor based on the guidelines provided in^42^, and used a stringent multiple testing significance threshold to account for the various number of PCs considered. Additional details are provided in **Supplementary Notes** and **Figures S1-2**.

### Contribution of taxa to covariance association signal

All the steps in the derivation of the statistical test are linear operations, which means that the contribution of features contributing to the MANOCCA association signal can be summed. Two types of contributions can be derived: the covariance contribution from each pair of taxa, and the sum of the covariance contribution assigned to each single taxon. The contribution of a given pair of taxa *i* and *j* to the covariance signal, *ϕ*(*P*_*ij*_), is defined as the square of the PCA loadings multiplied by the univariate association coefficient 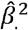 of the corresponding principal components with the considered predictor: 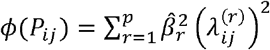, where 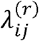 is the loading of the *ij* pair of taxa for PC *r*, and *p* is the total number of PCs included in the analysis. The single taxa contribution, *ψ*(*Y*_*i*_), can be derived by summing its contributions across all pairs: 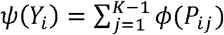, with *j* ≠ *i*, and *K* is the total number of pairs.

### Environmental association screening using MANOCCA

We applied MANOCCA to identify environmental factors associated with a change in covariances between taxa at the family, genus and species level. Milieu Interieur volunteers were asked to fill in a questionnaire of 44 pages, covering multiple panels such as demographics, lifestyle, and vaccination history. We selected the most relevant panels for the study, leading to the selection of 102 environmental factors. Among them was included diet information collected as part of the Nutrinet study^53^: the top three factors from the Nutrinet factors analysis, and the Nutrinet profiles were binarized to yes/no. We filtered out variables with more than half of the sample size in missing values, or a binary predictor with frequencies smaller than 5%. For categorical predictors displaying highly skewed distributions, outliers, defined as value three more standard deviation away from the mean were merged with a lower occurring category. A total of 80 environmental factors remained for analysis. After filtering, we ended up with a cohort of 938 individuals with complete shotgun sequencing, age, sex and body mass index (BMI) data. For the genus and family levels, we kept taxa abundant in at least 5% of the cohort, leading to a drop from 1,192 genera to 718 genera, and 216 families to 151 families. At the specie level, to avoid having too many species with regard to the sample size, we set the threshold to 40% of the cohort leading to a drop from 3,885 species to 675 species.

### Comparison with Manova and alpha diversity

For comparison purposes, we considered three alternative multivariate methods: a standard MANOVA and the alpha diversity, using the Shannon and Simpson indexes. The screening methodology was the same as the one used for MANOCCA, though some pre-processing adjustments were made to match the expected assumptions of each method. The MANOVA was applied to the taxa relative abundances from a given phylogenic level, which was processed following standards from the literature^54^: proportion followed by arcsin root transformation followed by a scaling. With **Y** the matrix of resulting taxa, *X* the considered predictor, and **C** a matrix of covariates. We applied the Wilk’s lambda test : **Y** ∼ *X* + **C**, and in more details with 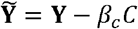 the residual matrix after adjustment from the covariates, 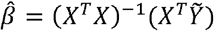 the regression coefficient and *N* the sample size, we could compute a *P*-value for the statistic :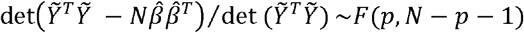 .

For the alpha diversity indexes, the raw abundances were used to the corresponding metric : 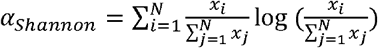and 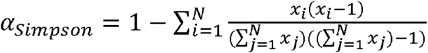 . The resulting α was tested in a standard univariate linear regression adjusted for the covariates: *α* ∼ *δ* _*X*_ *X*+ *δ*_*C*_*C*. The effect of *X* was assessed using a Wald test to the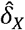.

### Deriving the covariance network

Networks of variation in the covariance were built using the top 1,000 co-abunding pairs derived using the MANOCCA test. In representing the network, we included three parameters: the total number of connections (qualified through the node size), the actual pairwise taxa connection (edges in the graph), and the direction of host factor effect on the covariance (decrease or increase of co-abundance). For each pair *Y*_*i*_*Y*_*j*_ adjusted for covariates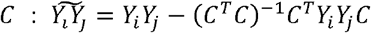, the direction of effect was derived using the sign of the regression coefficient for the predictor *X*: 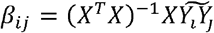 . For shared pairs with mixed direction of effect, the edge was colored in black. To facilitate the reading of the network, we coloured the node conditional on the association with each of the four predictors of interest, or shared among them. The ‘viridis’ cmap from *matplotlib* was used as colour scheme, with each shared taxa being a combination of original predictor colours.

### Comparison with other network-based approaches

For comparison purposes, we derived a permutation-based network inference approach for binary predictor, which we used to validate the MANOCCA network. In brief, we derive the pairwise covariance matrix for each categorical value, and then derive the empirical distribution of the correlation under the null by simulating *N*_*Permutations*_ covariances after shuffling the abundances of a bacterial taxa for each individual. Using a fixed detection threshold, we can then select pairs of taxa with extreme covariances. Since we are interested in variability of the covariance, we only keep the pairs uniquely detected across all values of the given predictor. When applied to compare results from the sex and smoking analyses, we ran 100,000 permutations, and retrieved the unique pairs detected in either group (women vs men, and non-smoker vs ever smoke). We also ran the SparCC^49^ correlation analysis, as this approach is commonly used and performed relatively well in a review of existing approaches^41^. We ran SparCC using the recommended parameters, deriving the *P*-values using 1,000 permutations, on both the simulated data (**Fig. S5**) and the real data (**Fig. S6**). Further descriptions of both approaches are provided in the **Supplementary Notes**.

### Using co-abundance for prediction purposes

We assessed the performances of a predictive model based on covariance across taxa. The implementation of a predictive model follows the standard used for multivariate linear model. For a given outcome *A* to be predicted, the estimated coefficients between *A* and 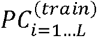 obtained in a training dataset from MANOCCA, 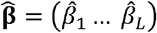 are projected on the principal component from an independent test dataset and summed up to form a predictive score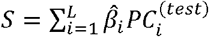 . Note that the dimensionality of the covariance data and the principal component analysis (PCA) step make the implementation slightly more complex. In particular, the principal components derived on the same variables for two independent samples might not always match, with structure in the data being capture by different components. To avoid this issue, PCA is not applied in the test data. Instead,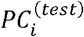 are derived by projecting out the loadings from the training sets: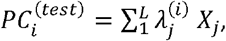, where 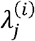 is the loading of variable 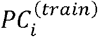 for product of taxa *i* obtained in the train data. It also implies that the test dataset should have the same dimension (i.e. approximately the same list of taxa) as the train dataset.

We applied this approach for the prediction of age, BMI, smoking and sex using taxa from the three lowest taxa levels (species, genus and family), using a 30-fold cross-validation, and without including other factors as covariate. For each of the 30 cross-validation, the dataset was randomly split into two independent sets: a training set including 90% of the data and a test set including the 10% remaining samples. We measured the accuracy of the predictive model using squared-correlation for continuous outcomes, derived as *cor* (*S,A*^*(test*)^ )^2^, and using the area under the receiver operating curve (AUC) for binary outcomes. The AUC is a common metric to quantify the predictive power of binary outcome. It equals the probability of correctly classifying a random sample from the test data.

## Data Availability

All data produced are available online at https://www.milieuinterieur.fr/en/

## Acknowledgment

This research was supported by the Agence Nationale pour la Recherche (ANR-20-CE15-0012-01). This work has been conducted as part of the INCEPTION program (Investissement d’Avenir grant ANR-16-CONV-0005). The Milieu Interieur consortium was also supported by the Agence Nationale pour la Recherche (ANR-10-LABX-69-01).

## Code availability

All code is available in Python and R at: https://gitlab.pasteur.fr/statistical-genetics/manocca

The complete network with full annotation is available in html at: https://gitlab.pasteur.fr/statistical-genetics/manocca

