## Supplements for "Gut microbiome co-abundance networks varies with age, sex, smoking status and body mass index"

**Supplementary Material**

### Supplementary Notes

#### Milieu Interieur gut microbiome data

Stool samples were collected during two visits. For participants that had two samples, we only kept the sample from the first visit. Shotgun metagenomic was applied to derive the bacterial profiles up to the specie level. Detailed description of the data generation can be found in Byrd et al^1^. In brief, stool specimens were collected in a double-lined sealable bag containing a GENbag Anaer atmosphere generator (Aerocult; Biomerieux). Then fresh samples were aliquoted into cryotubes and stored at −80°C. Stool aliquots were shipped to the CRO Diversigen for DNA extraction and shotgun metagenomic sequencing using an Illumina HiSeq 2500. In the end, 21 trillion raw paired-end reads from 1,359 samples from 946 of the donors were obtained. To process the reads, Illumina TruSeq adapters were trimmed with Trimmomatic v0.36^2^; low-quality and low-complexity reads were removed with prinseq-lite 0.20.4^3^; and Bowtie2 v2.1.0^4^ was used to remove reads mapping to PhiX or the PacBio human genome. After processing, there were on average 13.9 ± 2.9 million reads per sample. Of an initial 1,000 recruited donors, 44 were excluded from this analysis because of lack of consent for sharing their data outside of the MI consortium. An additional 10 donors were excluded because of technical issues in the extraction and sequencing steps (e.g., low DNA extraction yield), resulting in a sample size for the shotgun dataset of 946 donors. The final quantification was made using a Kraken-GTDB database^5^ on June 25, 2019. In the end, 23,505 genomes representing 13,446 unique bacterial species were downloaded and formatted into a Kraken2 database^6^.

#### Impact of dimension reduction in MANOCCA on association test.

When applied to taxa, we varied the number of principal components used in MANOCCA from two to one hundred but limited the number of PC analysed for each predictor based on the guidelines^7^, and used a stringent multiple testing significance threshold to account for the various number of PCs considered. The optimal number of principal components –corresponding to the smallest observed *P* value– varied substantially, but leans towards small values with a median of 11 PCs over the 80 variables analysed (**Fig. S1-S2**). However, it is substantially higher for three out of the four variables identified with maximum signal obtained at the genus level for 62, 83, 30 and 3 PCs for age, sex, smoking and BMI, respectively. This high number of PCs suggests that the association with the co-abundance involves a fairly large number of taxa.

#### Compositional data

Due to high-throughput sequencing (HTS) technologies, microbiome data is compositional in nature^8^. The counts given by 16S or shotgun sequencing depends on the sequencing depth and can vary substantially across samples. It can therefore not be related to the absolute count of bacteria in the input sample, meaning that only the relative abundance of microbial taxa can be used. This implies that every sample sums to a constant total. As pointed in previous studies^9,10^, this constraint can lead to issues such as spurious correlations, non-independence, and biased estimates when using traditional statistical methods. Typically, transformations like the centered log-ratio (CLR) transformation is utilized to address these challenges but brings other issues such as addressing 0 abundance counts^11^.

In this study, we applied our covariance-based test directly on the relative abundance. Although future work might investigate alternative metric, not modelling the compositional component should not impact the accuracy of our results. First, as showed in the simulation from **Figure S7**, the negative bias in correlation estimates discussed in Gloor et al^9^ appears when confronted with a small amount of OTUs in the model, but becomes negligible when reaching the sample size and number of OTU analyzed in the present study. Second, the proposed approach is not qualifying the correlation between taxa per se, but instead estimates changes in covariance conditional on a given variable of interest. This is a critical point. Indeed, even though some bias remains, it will only constitute a constant offset. Assuming the host factors considered in the present study are not associated with the total count of reads across individuals, this bias should have no impact on estimating the relationship between factors and taxa co-abundance.

#### Existing network-based approaches

There is no gold-standard method to investigate factors associated with taxa co-abundance, and existing methods are known to be sensitive to parameter tuning^12^. Existing approaches are typically based on the evaluation of networks co-occurrence and consists in three steps: i) estimating the pairwise correlation across taxa, ii) dichotomizing the correlation metric based on a given threshold in order to define a sparse network, and iii) comparing the inferred network based on descriptive statistics such the number of edges and nodes across various conditions. This approach is only applicable to binary (e.g. a disease status) or categorical predictors (e.g. multiple cohorts) with few categories, as it requires building a network for each group considered. Moreover, comparison between networks derived from different approaches is not straightforward, and typically rely on *ad hoc* empirical *P*-value derived through permutation, or test of heterogeneity applied at the taxa level^13^. Finally, not all methods allow for adjustment for potential covariates making the interpretation of the relative contribution of each factor difficult.

For comparison purposes, we used SparCC to identify the taxa displaying variability in co-abundance conditional on binary predictors. SparCC^14^ was developed to estimate correlations from high-throughput sequencing microbial communities data, and especially to address the compositional aspects of such data. It is based on the work from Aitchison^15^ on the estimation of the variance of the log ratio of two quantities of OTUs $i$ and $j$: $t_{ij}=Var[log\left( {x_{i}}/{x_{j}} \right)]$. Since $t_{ij}=Var[log{(x}_{i})]+Var[log{(x}_{j})]-2Cov[\log{(x}_{i}), \log\left( x_{j} \right)]$, the quantity $t_{ij}$ can be interpreted in relation to the basis abundance’s variances. SparCC relies on the assumption that the true correlation network is sparse and estimates the basis variance iteratively, computing all correlations and removing extreme correlations, which drives the sparse correlation assumption. Second, the log ratio cannot be derived for OUT that have an abundance of zero^11^. SparCC addresses this issue by applying a pseudo-count to all null occurrences of OTUs, assuming that all OTUs have a minimal occurrence in every sample. SparCC has proven very useful for the specific purpose of characterizing correlations. Its performance to identify predictors associated with variability in co-abundances is less clear^16^. In practice, an empirical *P*-value needs to be derived, which requires to run $N_{simu}$ times SparCC, which can be a strong computational burden and subject to noise if $N_{simu}$ is too small. Furthermore, SparCC can only be applied to categorical predictors, strongly limiting its applicability. Should the significant correlations be in the union, difference or intersection of correlations detected for each predictor values.

We also derived a naïve empirical permutation correlation test, which mimics an approach presented in a review of existing methods^12^. Here, we derived an original correlation value, and then shuffled the data $N_{s}$ times to derive an empirical distribution of correlations and derive an empirical *P*-value as $P={\min\left( \#above,\#below \right)}/{N_{s}}$. This approach displays severe limitations, the main one being the noise detections that requires an extreme number of permutations to eliminate.

In the case of MANOCCA, the usage of a linear framework allows to bypass any empirical simulation by using parametric statistics. Additionally, MANOCCA tests for a progressive change in correlations, allowing to take into account information from ordinal and continuous predictors. The resulting features are directly linked to changes in the covariance structure of the outcome. Finally, in comparison with the literature this approach allows to adjust for unwanted confounding variables in the model.

### Supplementary Figures

#### Figure S1. Co-abundance association signal conditional on the number of PCs used

Applying MANOCCA when the number of products is larger than the sample size requires reducing the dimension of the outcome data. Here we used principal component analysis (PCA) applied to the matrix of products, including only the top principal components (PCs) explaining the largest amount of total variance. We investigated the power of MANOCCA when varying the number of PCs kept in the model. The panels display the -log10(P) from the MANOCCA for each of the 80 predictors considered, as a function of the number of principal components selected, varying from 1 to 100 for the species (a), genus (b) and family (c) levels. The most associated predictors are highlighted in blue gradient.

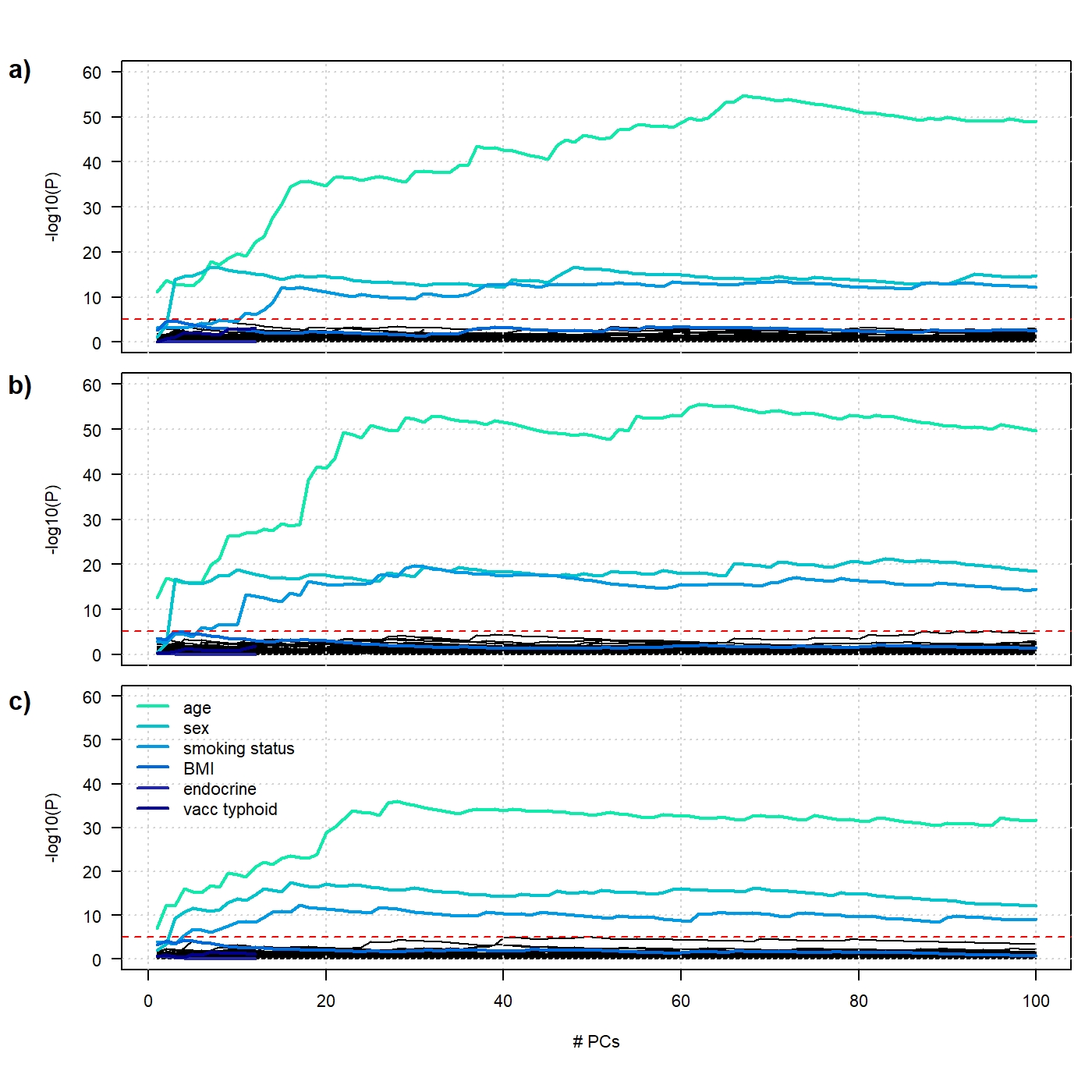

#### Figure S2. Optimal number of principal components per predictor

For each of the 80 environmental predictors considered, we recorded the best number of principal components (PCs) maximizing the association signal from MANOCCA. Results are presented for the species (dark red), genus (red) and family (pink) levels. At the bottom we recorded for each taxonomic level the median of the number of PCs kept across all predictors.

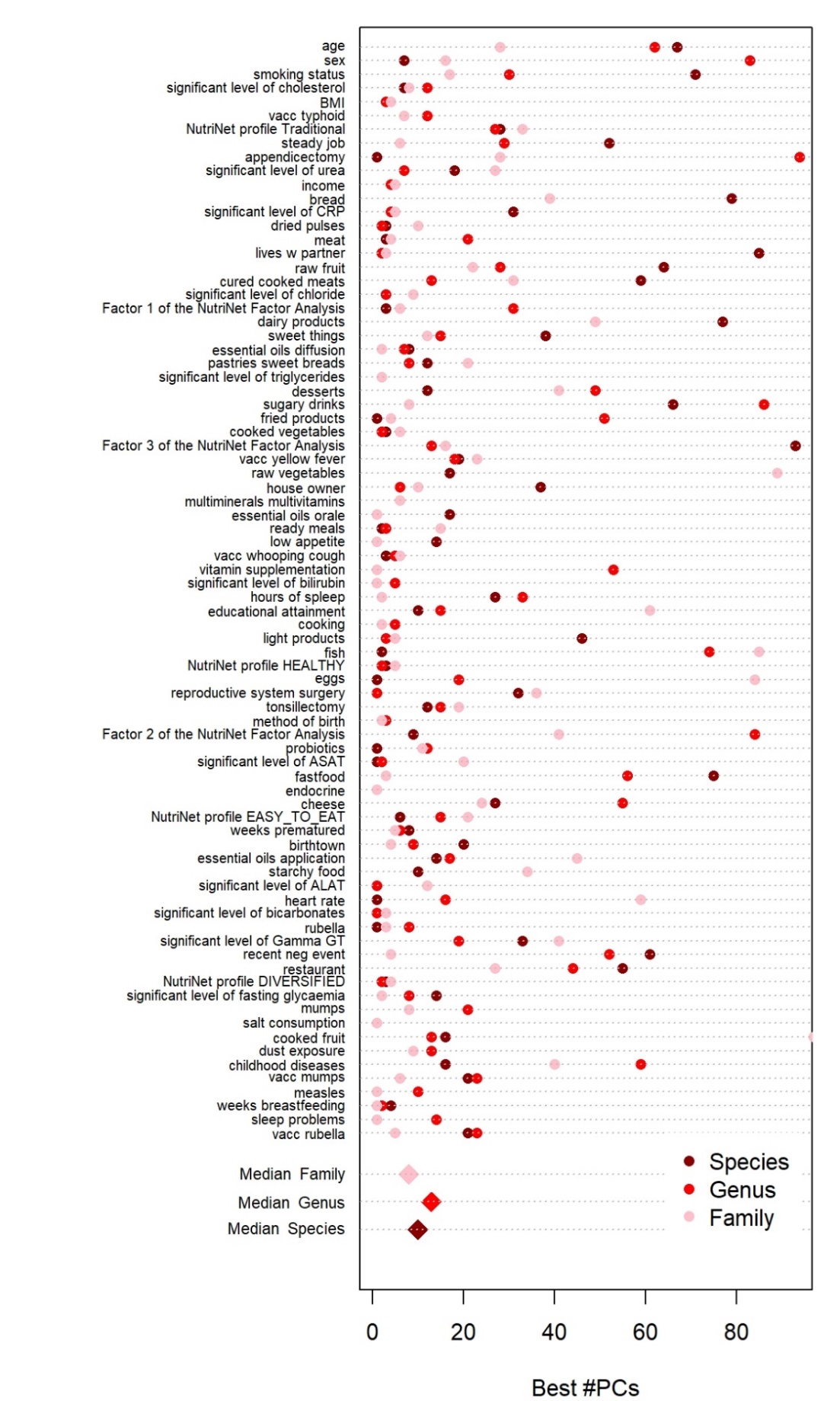

#### Figure S3. Sensitivity analysis: signal for sex after random subsampling

To assess the sensitivity of the co-abundance signal observed, we re-run our analysis on random subsets of 20 to 700 genus taxa (from a total of 718 genera). For each sample size, we sampled 100 subsets and derived the *P*-value from MANOCCA. The four panels show boxplot across the series of 100 subsets for the four host factors of interest (age, sex, BMI, and smoking).

=
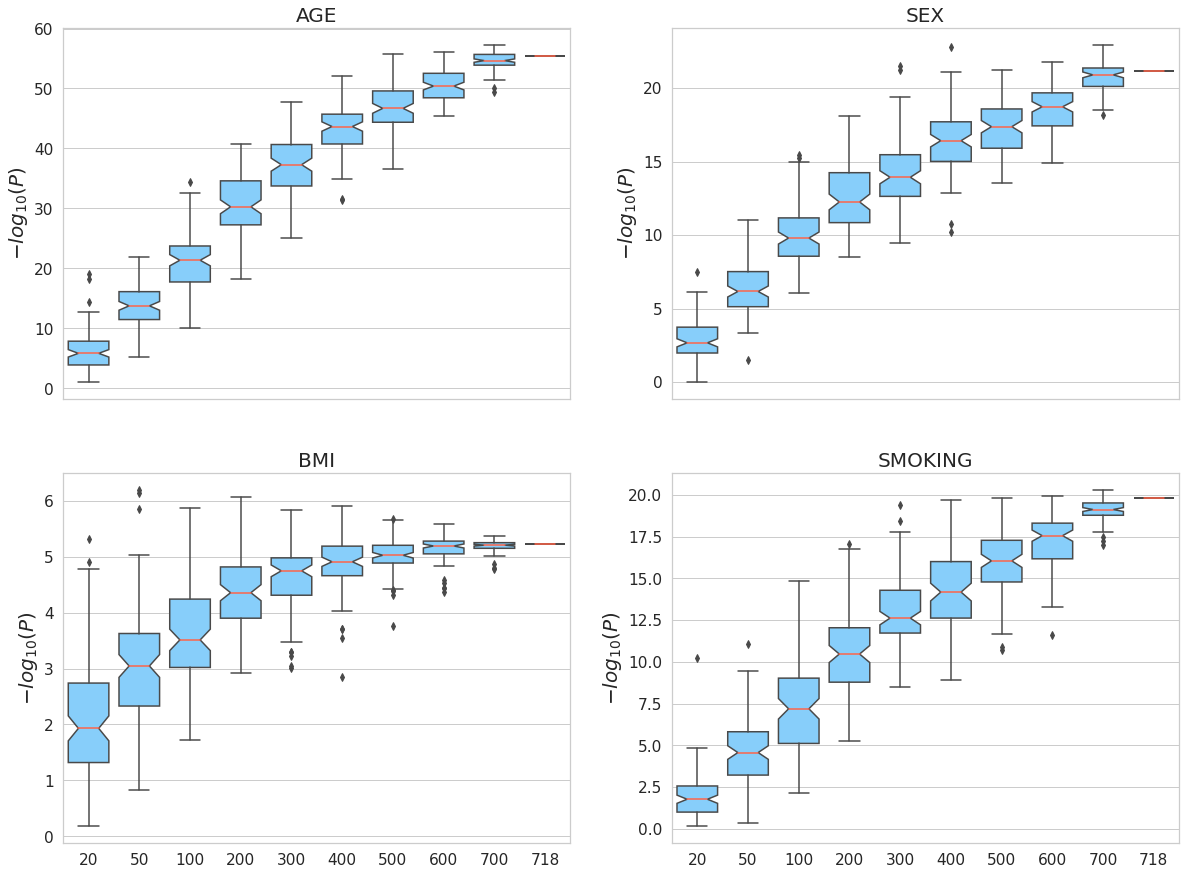

#### Figure S4. Covariance versus mean effect

We derived the contribution to the covariance signal from each taxon for the top four associated host variables: age, sex, BMI and smoking status. We compared those contributions to a univariate screening of mean abundance for the same taxa. Panel a) displays the normalized contribution across all the 721 genus taxa for MANOCCA. Panel b) displays the distribution of the contribution (blue), as compared to a null distribution obtained after shuffling the predictor considered (grey). Panel c) displays the -log10(P-value) from the univariate linear regression for the same taxa. The dotted blue line displays the Bonferroni threshold for the univariate tests.

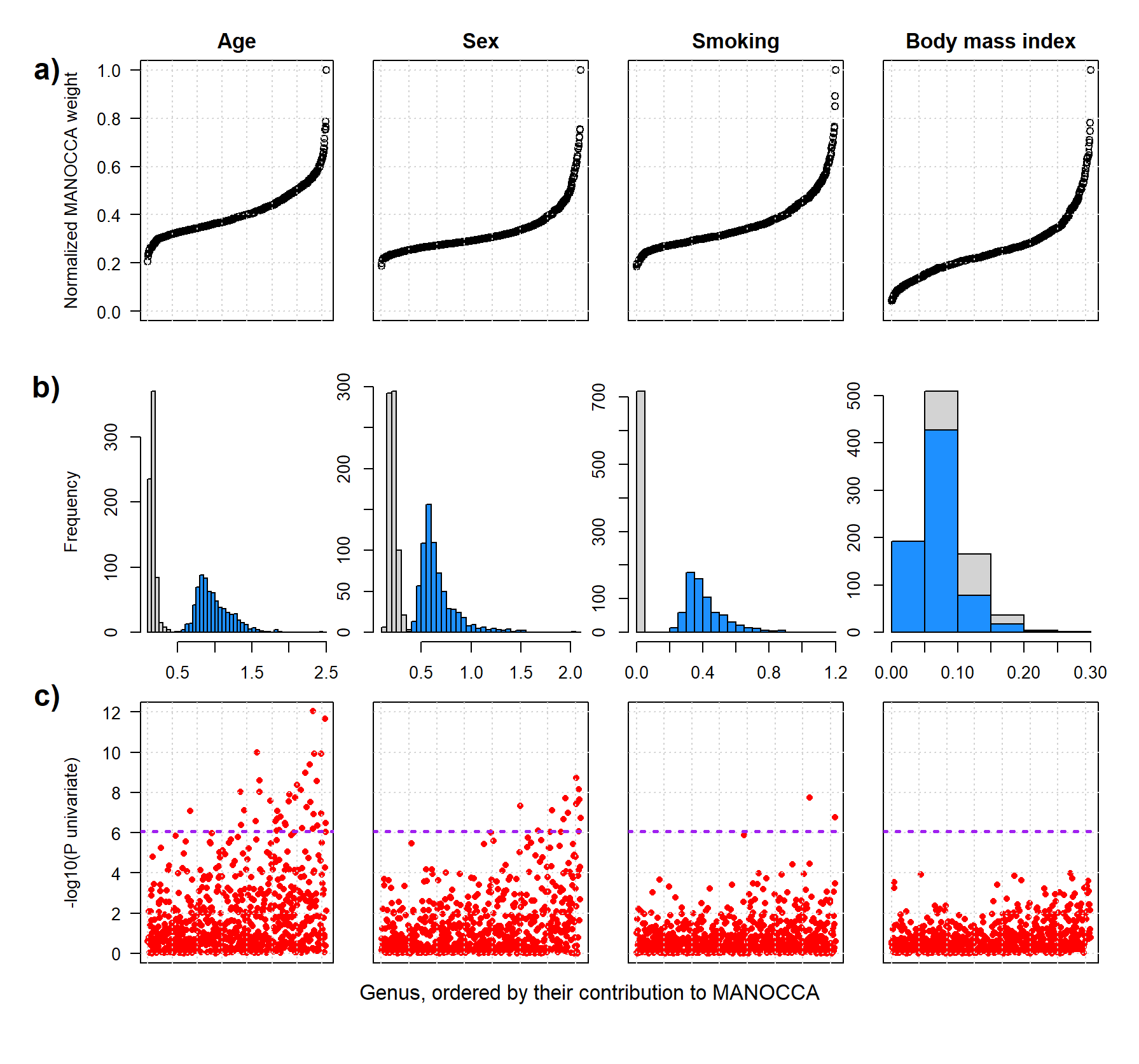

#### Figure S5. Methods comparison on simulated data

We ran a simulation study where data where 100 variables were drawn two multivariate normal distribution with correlation matrices $C_{1}$ and $C_{2}$, respectively. Correlation matrix $C_{1}$ was simulated from random noise, and the second correlation matrix $C_{2}$ was a copy of$C_{1}$ but the upper left square of 20 variables was modified to induce strong correlations. Both matrices are presented in the lower and upper triangles of panel a), respectively. We generated a dataset including 400 samples drawn using $C_{1}$, and 600 samples drawn using$C_{2}$. We ran a permutation-based approach, SparCC and MANOCCA on the simulated dataset. Panel b) and c) show in the lower triangle the $-log10(Pvalue)$ for each cell on data generated using $C_{1}$ and using $C_{2}$ in the upper triangle. Panel d) displays the top 190 normalized MANOCCA features derived using the simulated data as outcome and the binary predictor 0 if data was generated using $C_{1}$ and 1 if using $C_{2}$.

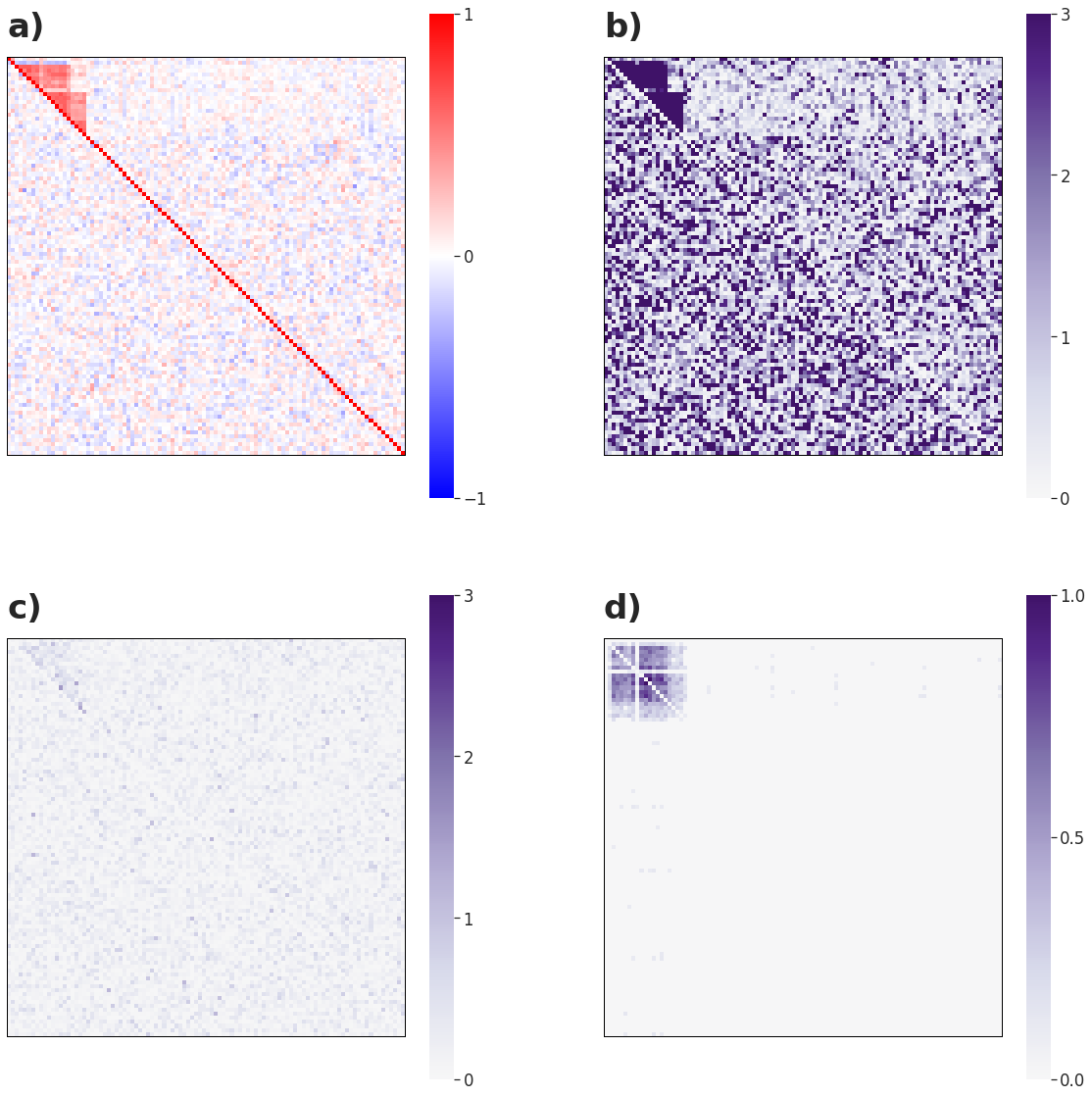

#### Figure S6. Comparison with existing network approaches

We estimated the significance of the overlap between networks of impacted co-abundances derived using the MANOCCA’s contributing products, and two alternative approaches, an *ad hoc* permutation-based approach and SparCC. Because the two latter approaches are only applicable to binary predictor, we conducted our comparison only on sex (panel a)) and using a binary version of smoking (never/ever smoking) (panel b)). We applied all methods at the genus level, comparing a total of 257,403 pairs of taxa. Pairs were ranked based on their MANOCCA’s contribution, and the *P*-value for the permutation-based approach and SparCC. Pairs of genera belonging to the top percentiles, varied from 0.01 to 0.9, were extracted for each approach, and the significance of the overlap between the two set was derived using a Binomial test. For both predictors and both approaches, the most significant overlap was observed when using the top 50% contributing pairs of MANOCCA and the top 10% associated pairs from the permutation-based approach.

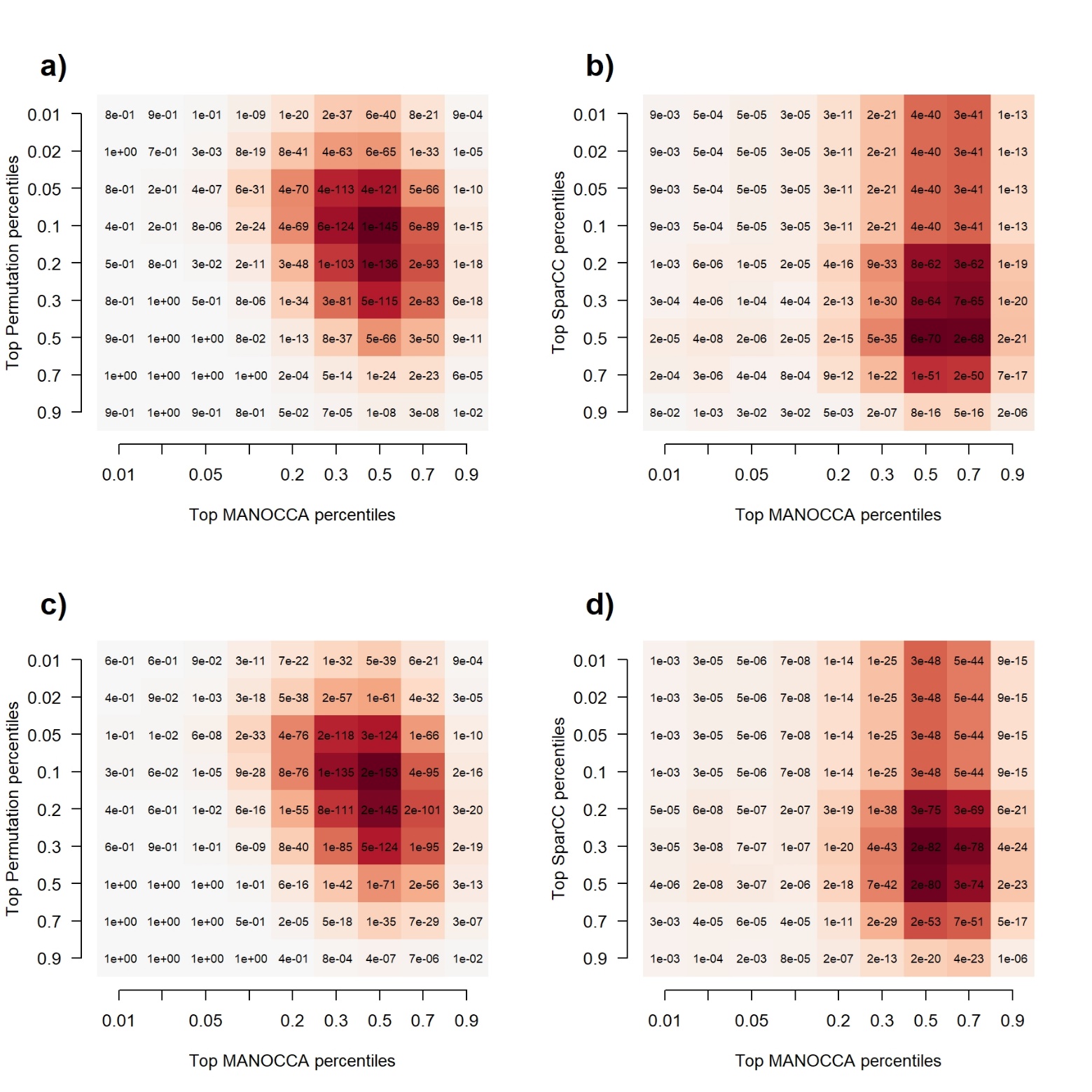

#### Figure S7. Evolution of compositional issue with regards to the number of variables

We assessed the extent of spurious correlations due to the compositional component of microbiome data across series of 50 replicates with various sample size and number of variables. We considered eight settings with [N,M], where N is the sample size and M the number of variables: [20,10] (M1), [50,20] (M2), [100,30](M3), [200,40](M4), [400,60](M5), [600,100](M6), [800,300](M7), [900,700](M8). For each setting we derived the mean difference between correlations computed on the whole dataset and correlations computed on a subset of 1/2, 3/4, 9/10 of the variables. In grey we display the basis expected randomness of correlations between two normal distributions with no compositional aspect. Panels a-c) display results with data simulated using a uniform distribution between 10 and 10000 and for increase proportions of subsets. Panel d-f) display results on the real data using the 718 genera.

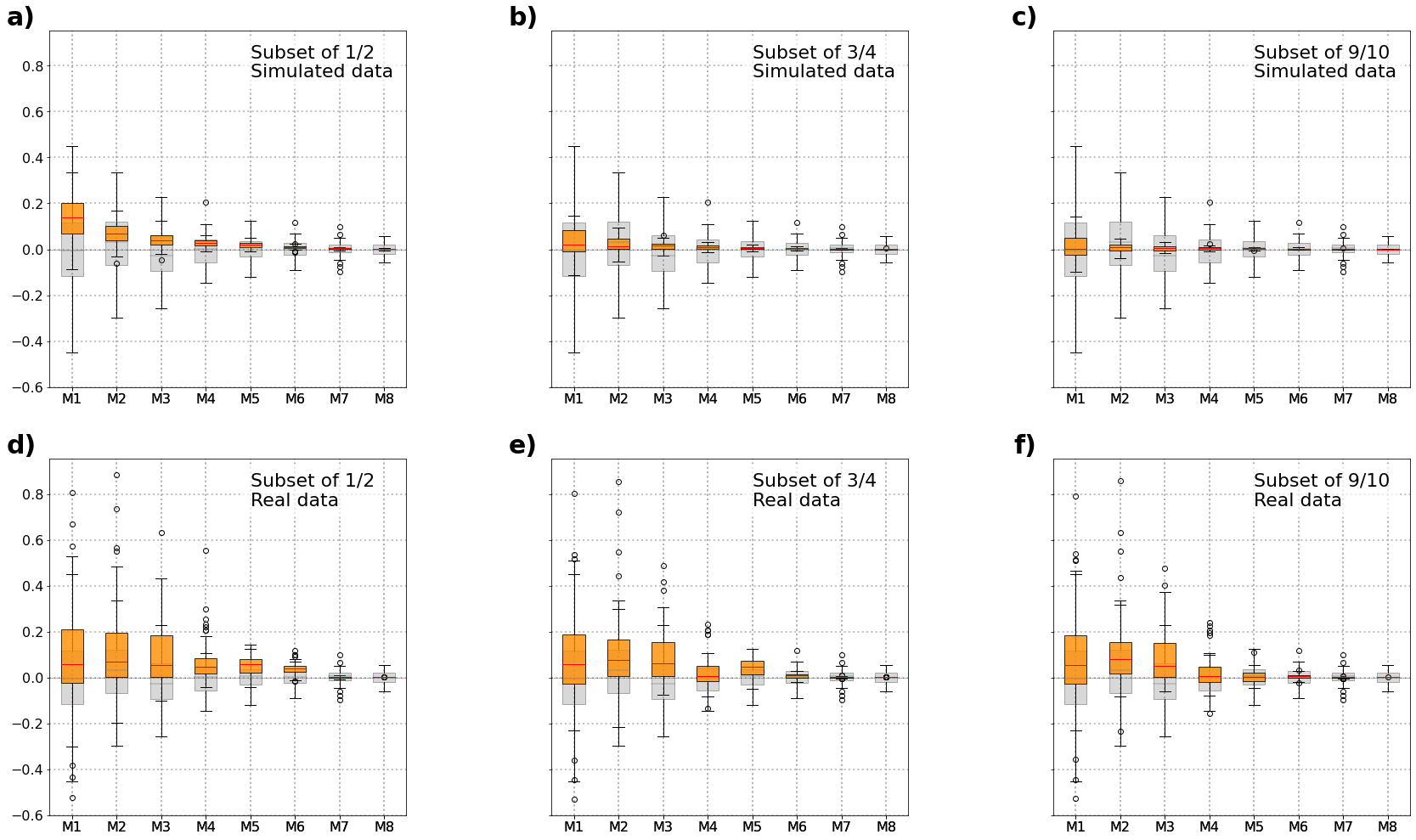
